# Public Perception of COVID-19 Vaccines through Analysis of Twitter Content and Users

**DOI:** 10.1101/2021.04.19.21255701

**Authors:** Sameh N. Saleh, Samuel A. McDonald, Mujeeb A. Basit, Sanat Kumar, Reuben J. Arasaratnam, Trish M. Perl, Christoph U. Lehmann, Richard J. Medford

## Abstract

Twitter is a robust medium to understand wide-scale, organic public perception about the COVID-19 vaccine. In this cross-sectional observational study, we evaluated 2.4 million English tweets from nearly 1 million user accounts matching keywords ((‘covid*’ OR ‘coronavirus’) AND ‘vaccine’) during vaccine development from Feb 1^st^ through Dec 11^th^, 2020. We applied topic modeling, sentiment and emotion analysis, and demographic inference of users on the COVID-19 vaccine related tweets to provide insight into the evolution of public attitudes. Individuals generated 87.9% (n=834,224) of tweets. Of individuals, men (n=560,824) outnumbered women (n=273,400) by 2:1 and 39.5% (n=329,776) of individuals were ≥ 40 years old. Daily mean sentiment fluctuated congruent with news events, but overall trended positively. Trust, anticipation, and fear were the three most predominant emotions; while fear was the most predominant emotion early in the study period, trust outpaced fear from April 2020 onward. Fear was more prevalent in tweets by individuals (26.3% vs. organizations 19.4%; p<0.001), specifically among women (28.4% vs. males 25.4%; p <0.001). Multiple topics had a monthly trend towards more positive sentiment. Tweets comparing COVID-19 to the influenza vaccine had strongly negative early sentiment but improved over time. Our findings are concerning for COVID-19 vaccine hesitancy, but also identify targets for educational interventions.

## Introduction

With the global continuation of the COVID-19 pandemic, the large-scale administration of a SARS-CoV-2 vaccine (referred from here on as the COVID-19 vaccine) is crucial to achieve herd immunity and curtail further spread of the virus^1^. As governments work to approve and distribute safe and effective vaccines,^2^ important questions regarding vaccination willingness persist: What are the attitudes and perceptions of the public^3^ to these vaccines and how can they affect vaccine uptake^4^? These questions are important to develop an education and outreach approach to achieve the desired vaccine penetration to achieve herd immunity^5^. In 2019, prior to the COVID-19 pandemic, the World Health Organization (WHO) had identified vaccine hesitancy as one of the top 10 greatest global health threats^6^. While surveys on attitudes and perception of a COVID-19 vaccine show significant vaccine hesitancy among the general population^7–9^ and health care providers^10,11^, studies remain small in size, tend to focus on local participants, are prone to sampling error from non-probability sampling and reporting bias, and perhaps most poignantly, cannot capture real-time changes in vaccine willingness. Crowdfunding platforms may provide an indication of emerging community needs related to COVID-19 but fail to provide a continuous assessment of community sentiment^12^.

Twitter, the microblogging platform, with over 187 million daily monetizable active users,^13^ serves as a robust medium to better understand wide-scale, organic public perception about the COVID-19 vaccine. With nearly 400 million mentions, #COVID19 was the most used hashtag on Twitter in 2020^14^. Social media has become increasingly recognized for its rapid information dissemination (whether accurate or not) and dispersion of sentiment that quickly crosses geographic and social boundaries^15^. Analysis of social media text can inform real-time changes and evolution in population-level attitudes^16^. As evident with the rise of the “infodemic” during the COVID-19 pandemic, Twitter has become a particularly useful data source in public health and healthcare-related research^17^ and has been repeatedly used to study public sentiment and understand trends throughout the COVID-19 pandemic^18–21^. Earlier in the COVID-19 pandemic, we were able to demonstrate initial public sentiment regarding the virus, its origin and spread, and measures to limit its spread^22^ as well as early support for social distancing^23^ on Twitter.

Social media, and specifically Twitter, has been shown to be a major factor in vaccine uptake and should be monitored and potentially used for interventions to address vaccine hesitancy^24^. Examining sentiments towards the influenza A H1N1 vaccine in 2009 showed that projected vaccination rates based on Twitter sentiment were similar to vaccination rates estimated by traditional phone surveys used by the Centers for Disease Control and Prevention (CDC)^25^. A previous study noted information exposure on Twitter may account for differences in human papillomavirus (HPV) vaccine uptake that are not accounted for by socioeconomic factors like education, insurance, or poverty^26^. Another study noted that there is a significant relationship between social media use by the public and organized action and public doubts of vaccine safety^27^.

We aimed to apply content and sentiment analysis on COVID-19 vaccine related tweets as well as analysis of the responsible, originating user accounts to provide insight into the evolution of public attitudes about the COVID-19 vaccines over time. We hypothesized that content analysis from the start of the pandemic will identify important themes of discussion (especially those with negative sentiment or evidence of misinformation) throughout the vaccine development process that would inform health care officials, public health agencies, and policy makers and could be used to aid in the outreach and educational interventions for the COVID-19 vaccine to the general public.

## Methods

### Data Source

We performed a cross-sectional observational study of English-language tweets obtained by matching the keywords ((‘covid*’ OR ‘coronavirus’) AND ‘vaccine’) from February 1, 2020 to December 11th, 2020. December 11th was chosen as an end date to mark the United States Food and Drug Administration’s first emergency use authorization of a COVID-19 vaccine^28^. We used the *snscrape* library^29^ to obtain (“scrape”) tweets identified through Twitter’s advanced search tool, which returns a relevant sample of tweets. We manually reviewed a random subsample of 1,000 tweets and verified the tweets’ relevance to the topic of COVID-19 vaccination. We extracted 21 and 20 variables related to the tweets and to the posting user accounts, respectively (Supplemental Table S1 and S2).

### Data Processing

We measured total daily tweets and completed descriptive statistics for collected variables. We applied natural language processing techniques to process, analyze, and visualize the text from tweets. To preprocess the tweet text for analysis (“cleaning”), we removed hyperlinks, user tags, and words of little analytical value. We also returned words to their root form and segmented text into oneN- and two-word terms. Further details are discussed in Supplemental Appendix A. We visualized the top 300 processed terms as a word cloud with larger font size representing greater term frequency. All analyses were conducted using Python, version 3.8.2 (Python Software foundation). Institutional review board approval was not required because this study used only publicly available data.

### Sentiment Analysis

Sentiment analysis describes the affect of a piece of text — the intrinsic attractiveness or aversiveness of a subject such as events, objects, or situations^30^. We used the Valence Aware Dictionary and sEntiment Reasoner (VADER)^31^ to analyze the sentiment polarity of a tweet. VADER, a lexicon and rule-based tool, was particularly designed for sentiment analysis for social media text. In addition to regular words, VADER leverages punctuation, emoticons, emojis, sentiment-laden slang words and acronyms, as well as syntax and capitalization schemas to inform labeling of a positive, neutral, and negative score for each document. These three scores were combined to form a normalized, weighted composite score. Overall positive (≥-0.05), neutral (−0.05 to 0.05), and negative (≤-0.05) sentiments are defined at standardized composite score thresholds. When sentiment has been aggregated, we refer to an average sentiment of ≥-0.05 as positive and ≤-0.05 as negative. Trends in sentiment over time were determined using the Mann-Kendall trend test. We used the *TextBlob* library^32^ to label each tweet from a range of 0 (objective) to 1 (subjective) where objective tweets relay factual information and subjective tweets typically communicate an opinion or belief. Finally, we used the *NRCLex* library to label words within each tweet with corresponding emotional affects (i.e., Plutchik’s wheel of emotions which include anger, anticipation, fear, disgust, joy, sadness, surprise, and trust) based on the National Research Council Canada (NRC) affect lexicon^33^. Based on these labels, we identified tweets with their primary emotion and visualized how the proportion of eligible tweets (i.e., those with an identified primary emotion) with a particular primary emotion changed over time.

### Topic Modeling

After cleaning the tweets to distill analyzable text as described in the methods, we applied a machine learning algorithm called Correlation Explanation (CorEx)^34^ to identify clusters of topics for all tweets. CorEx identifies the most informative topics based on a set of latent factors that best explain the correlations in the data in turn maximizing the total correlation or the multivariate mutual information^35^. Each document (in our case, each tweet) may include multiple topics. We iterated through a range from 2 to 20 topics and trained a separate model for each number of topics with the goal of identifying the model with the maximum total correlation, ultimately choosing 15 topics for the topic model. We presented the top 20 words for each topic cluster to author CUL without prior access to individual tweets from the dataset to manually label a theme for each topic. The manually labeled topic labels were reviewed by two other authors SNS and RJM with unanimous agreement. We visualized the monthly distribution of topics over time and utilized a heat map to visualize how the mean sentiment of each topic has changed per month.

### User Exploration and Demographic Inference

Given that each tweet has one authoring account, we identified all unique user accounts in our dataset and provided descriptive statistics with metadata available for the users, including the launch date of the account, followers (accounts following them), follows (accounts they follow), lifetime posts, likes, and media shared, as well as profile pictures, description information, and verified status (badge to indicate an account of public interest that has been verified to be authentic). To better understand demographic differences, we applied a previously validated deep learning system through the *m3inference* library^36^ to infer the account user as an individual or an organization and if labeled as an individual, their gender (female or male) and age group (≤18 years old, 19-29 years old, 30-39 years old, and ≥ 40 years old) based on multimodal input that includes username, display name, description, and profile picture image. Each label using the algorithm has an accompanying probability. The automatic demographic detection was particularly designed for Twitter profiles for health-related cohort studies^37^. We provided summary statistics for the demographics identified and stratified sentiment and subjectivity analyses by the different demographic groups to evaluate for differences. We used Mann-Whitney U and χ^2^ where appropriate to determine significance. Alpha level of significance was set a priori at 0.05 and all hypothesis testing was two-sided. We did not adjust for multiple comparisons as this was an exploratory study and should be interpreted as hypothesis-generating.

## Results

A total of 2,356,285 tweets were extracted for the study period, of which 2,287,344 tweets were English-only and included for evaluation. The tweets were generated by 948,666 accounts which had been active for an average of 6.9 years (interquartile range [IQR], 2.6 - 10.0) with a median of 267 (IQR, 55 - 1,100) followers and 3,600 (519 - 15,572) lifetime likes. Only 2.9% (n=27,443) of accounts were verified (Table 1). Of the tweets analyzed, 54% (n=1,235,575) had a link, 40.1% (n=916,585) mentioned other twitter accounts, 18.1% (n=414,173) used hashtags, and 11.9% (n=273,278) contained media like an image or video. In terms of engagement, 41.3% (n=943,639), 24.0% (n=548,863), and 20.7% (n=473,204) of tweets received likes, replies, and retweets, respectively (Table 1). Individuals (vs. organizations) generated 87.9% (n=834,224) of tweets. Of individuals, men (n=560,824) outnumbered women (n=273,400) by 2:1 and 39.5% (n = 329,776) of individuals were ≥ 40 years old (Table 1).

**Table 1.**
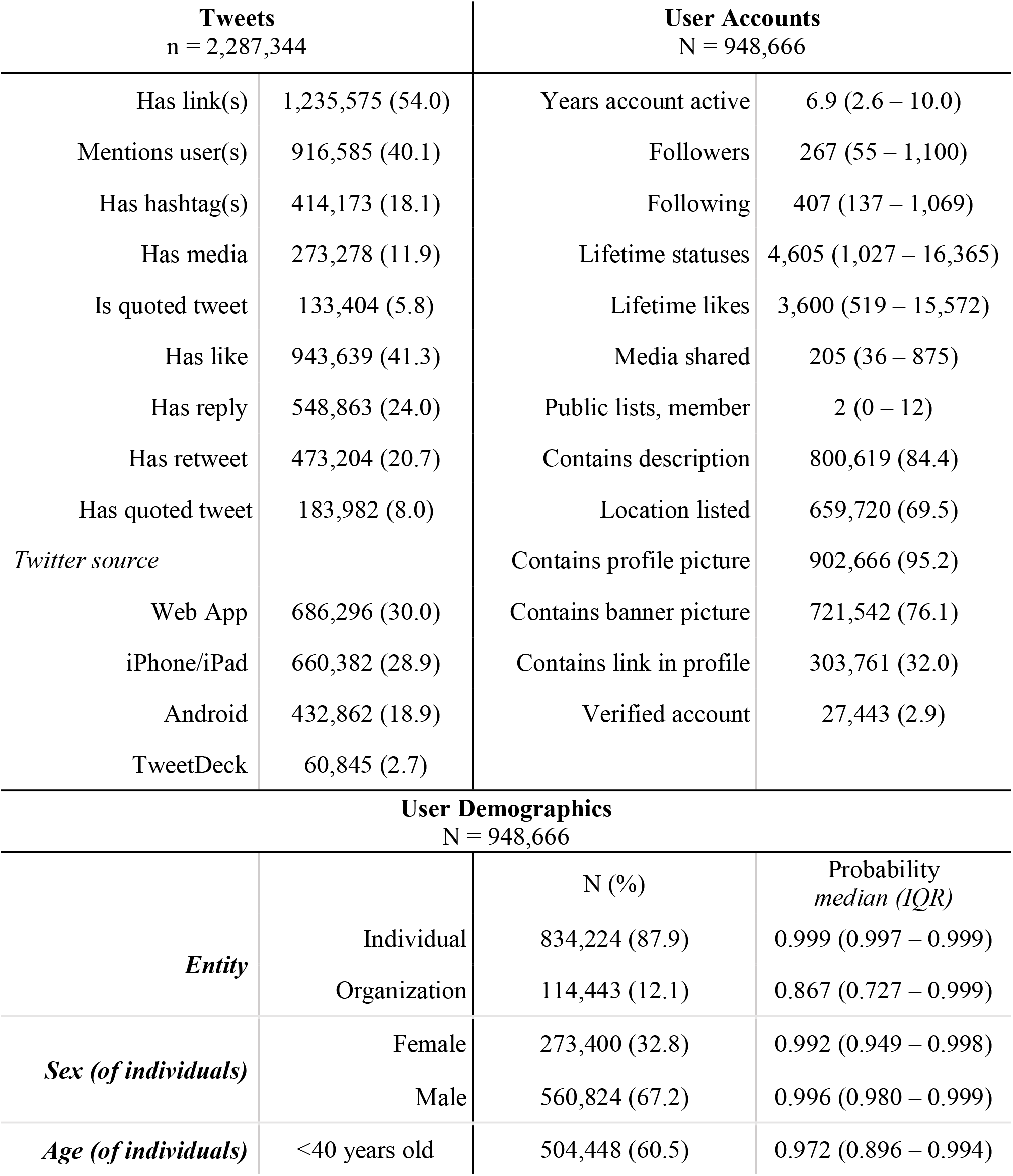
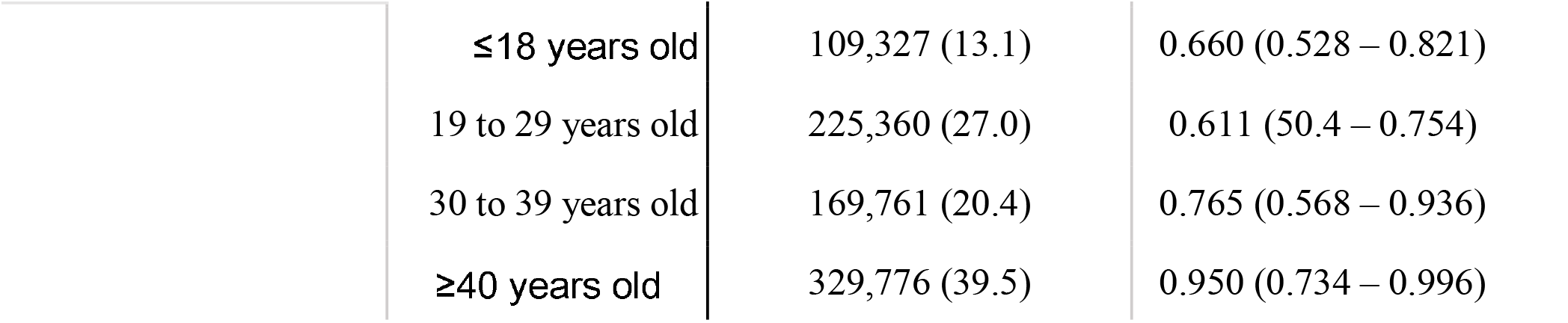
Tweet and user account characteristics are shown on top and inferred user demographics are shown on bottom.

Daily tweets abruptly spiked to 51,176 tweets on November 9^th^, the day Pfizer and BioNTech announced their vaccine’s effectiveness^38^ (up from 4,052 tweets on November 8^th^) and peaked on December 8^th^ with 55,779 tweets. Tweets from November 1^st^ to the end of the study period on December 11th accounted for 39.8% (n = 910,593) of all tweets (Figure S1). The corpus of tweets contained over 62.5 million words and 416 million characters. The ten most commonly tweeted terms and their frequencies were as follows: “people” (228,482), “trial” (206,310), “take” (181,598), “flu” (159,043), “trump” (149,042), “first” (147,103), “make” (142,242), “test” (131,719), “need” (126,846), and “one” (122,966). Figure 1 displays a word cloud of the top 300 words with larger font size concordant with frequency.

**Figure 1.**
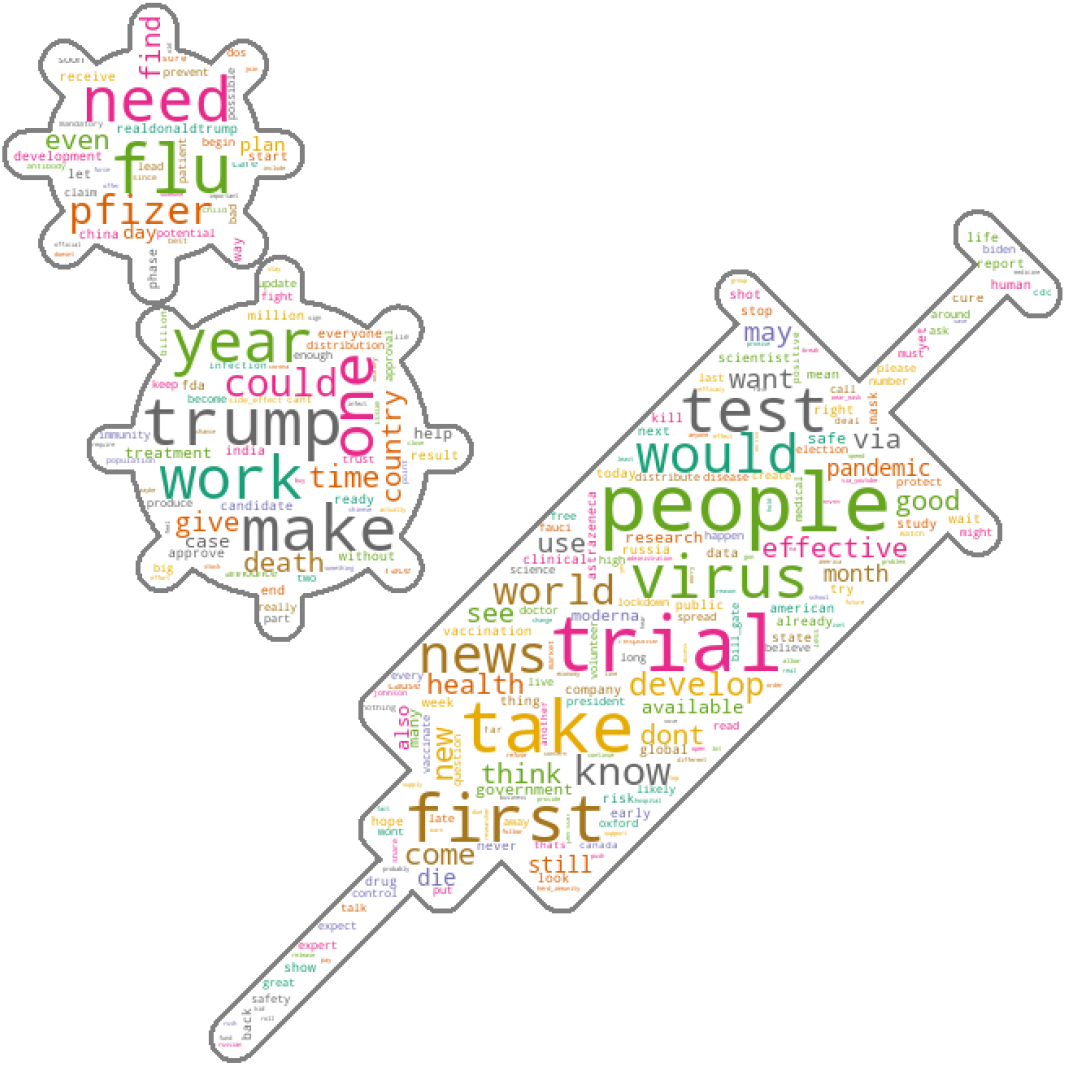
Word cloud of top 300 words related to COVID-19 and vaccine. Larger fonts represent higher frequency in the corpus after preprocessing text.

Daily mean sentiment of tweets fluctuated congruent with news events, but overall trended positively throughout the study period (Mann-Kendall statistic=10,122; tau=0.218; p<0.001) (Figure 2a). Several days in early to mid-March and on October 13th saw particularly negative sentiments, coinciding with news of the declaration of a pandemic by the WHO and Johnson & Johnson’s halting of their vaccine trial on October 12th^39^, respectively. Highest daily mean positive sentiment revolved around Moderna’s July 14th announcement of a safe vaccine with “robust immune response” in an early trial^40^ and Pfizer’s November 9th announcement of over 90% effectiveness of its vaccine^38^. Twitter accounts representing organizations had more positive sentiments than tweets from individuals (median weekly difference, 0.118; IQR, 0.091 to 0.144), but there was no significant difference in polarity for age (median weekly difference, 0.006; IQR, -0.011 to 0.019) and only minimal positive difference for males (median weekly difference, 0.030; IQR, 0.012 to 0.044) (Figure 2b-d).

**Figure 2a-d.**
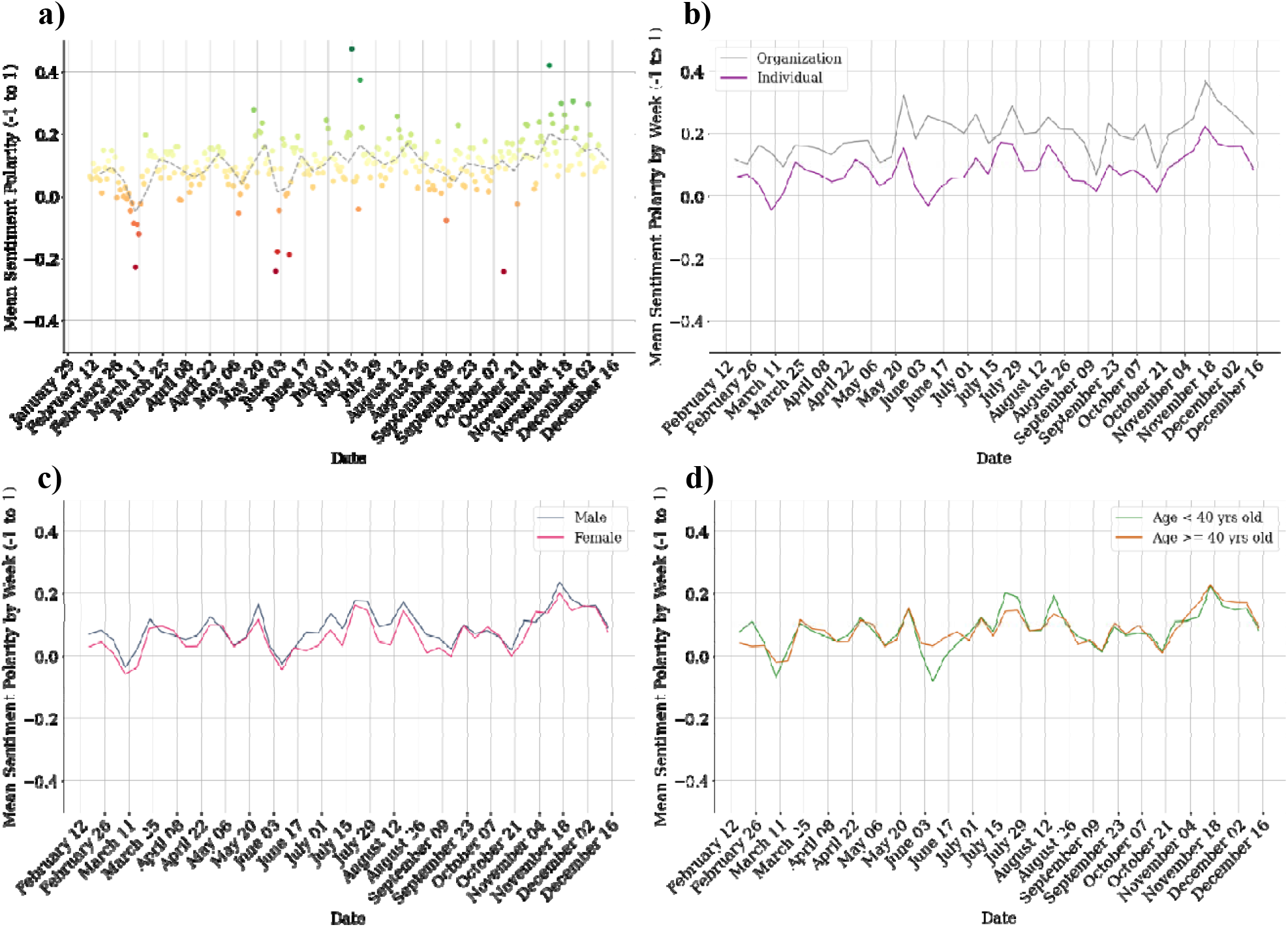
Mean sentiment polarity shown by day (as points) and by week (as a dashed line). Each tweet was labeled as primarily negative (−1), neutral (0), or positive (1). **b)** Mean weekly polarity stratified by individual versus organization. **c)** Mean weekly polarity stratified by gender for individual accounts. **d)** Mean weekly polarity stratified by age more or less than 40 years than for individual accounts.

The sentiment trends were reflected by the primary emotions identified in the COVID-19 vaccine tweets by month (Figure 3a). Fear started as the most prevalent primary emotion in nearly 40% of eligible tweets early on but decreased to under 20% by the end of the study period. Conversely, trust increased from below 20% to around 40% and outpaced fear in April 2020, maintaining as the most prevalent primary emotion thereafter. Anticipation was the second most prevalent primary emotion for most of the study period, steadily ranging from 25% to 30%. All other emotions were consistently expressed as the predominant emotion in less than 10% of eligible tweets. Individuals had an increased predominance of fear (26.3% vs. 19.4%; p<0.001) and decreased predominance of anticipation (25.9% vs. 33.6%; p<0.001) and trust (32.5% vs. 35.2%; p<0.001). For individual accounts, women had more fear (28.4% vs. males 25.4%; p<0.001) with less anticipation (23.8% vs. 26.8%; p<0.001) than men, but no significant difference in trust (32.3% vs. 32.5%, p=0.11). Those less than 40 years old had more fear (26.6% vs. 26.0%; p<0.001) and less trust (32.0% vs. 33.0%; p<0.001) (Figure 3b-d). Tweets throughout the year tended to be more objective (where 0 is fully objective and 1 as fully subjective) with limited daily variation (overall mean 0.359; std 0.028) (Figure S2).

**Figure 3a-d.**
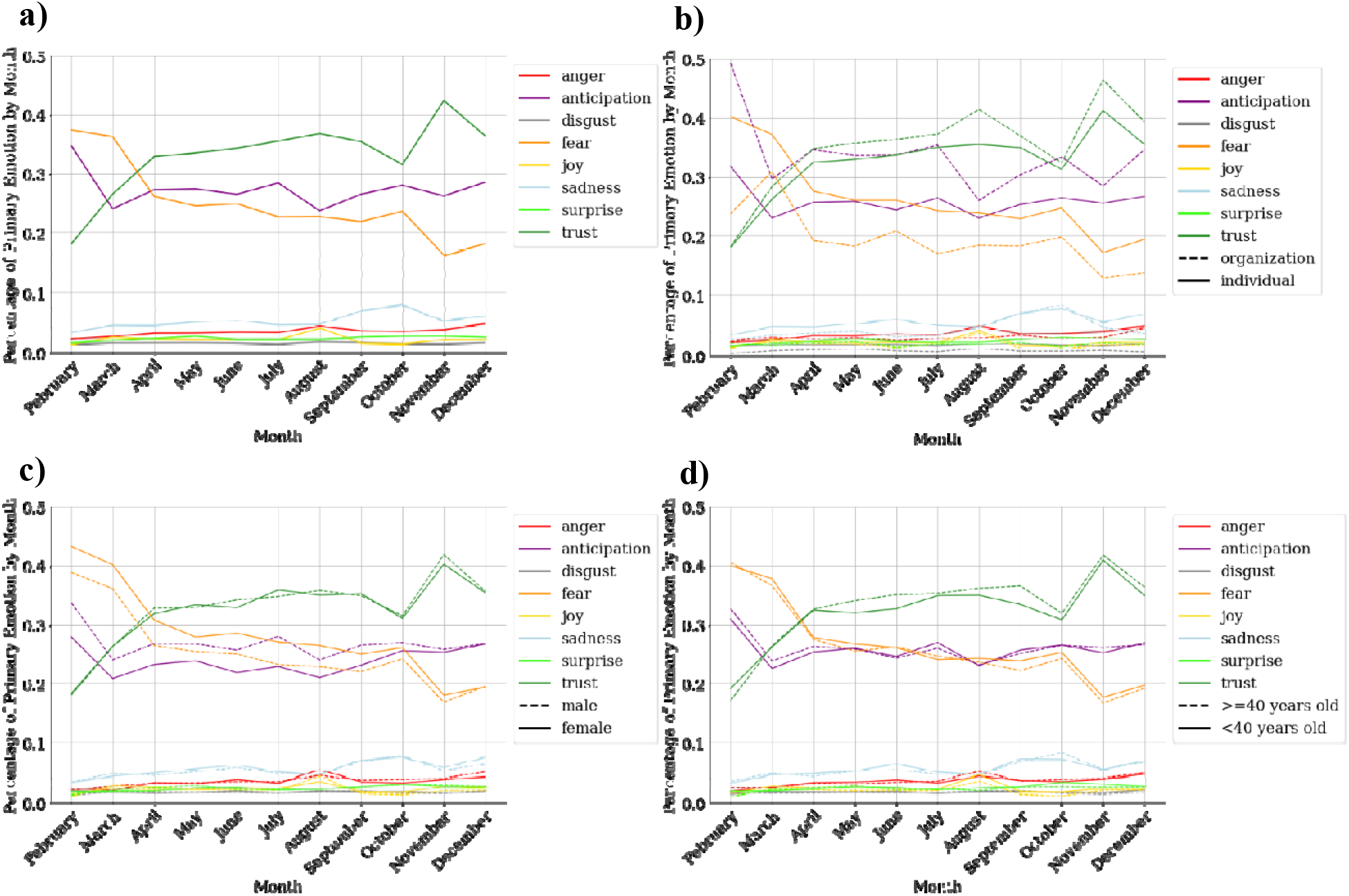
Percent of tweets with primary emotion per month **a)** overall, **b)** stratified by individual versus organization, **c)** stratified by gender for individual accounts, and **d)** stratified by age more or less than 40 years than for individual accounts. Only tweets with a predominant primary emotion (n = 1,489,027) are included.

Table 2 shows each topic label with their key words and sample tweets. Figure S3 shows the 15 topics obtained from topic modeling with the proportion of tweets per month that contained each topic. The dominant topic (topic 15) focused on mask use and public reactions. Discussions about misinformation and conspiracy theories comprised the next most common topic, peaking in May and staying relatively consistent from July through December. Tweets related to the Indian and Russian governments’ decision on producing and using the Sputnik V vaccine (topic 2) spiked in August. Discussion of Emergency Use Authorizations (EUA) and vaccine approvals (topic 12) did not spike until November 2020 with the approval of the Pfizer and Moderna vaccines. Several topics had strong mean positive sentiments throughout the study period, including discussions of biotechnology companies and the stock market (topic 3), vaccination firsts (topic 4), vaccine development (topic 6), and EUAs (topic 12). Other topics showed a progressive trend from positive to negative throughout the study period including discussion of US politics and the election (topic 1), the FDA and CDC (topic 14), and mask use and public reactions (topic 15). Tweets comparing COVID-19 to influenza (topic 5) and its vaccine had strongly negative early sentiment but improved over time (Figure 4). Compared to the rest of individual users (n=810,318), those exhibiting negative sentiment posting about topic 5 (n=51,686) were proportionally more likely to be ≥ 40 years old (45.1% vs. 39.6%; p <0.001) and female (34.0% vs. 32.7%; p <0.001). The only other topic with persistently negative sentiment was discussion of misinformation and conspiracy theories (topic 13). Those exhibiting negative sentiment posting about topic 13 (n=166,819) were more likely than other user accounts (n=741,388) to be individuals (90.9% vs. 87.3%; p <0.001) and of those individual accounts, more likely to be female (34.4% vs. 32.4%; p<0.001).

**Table 2.**
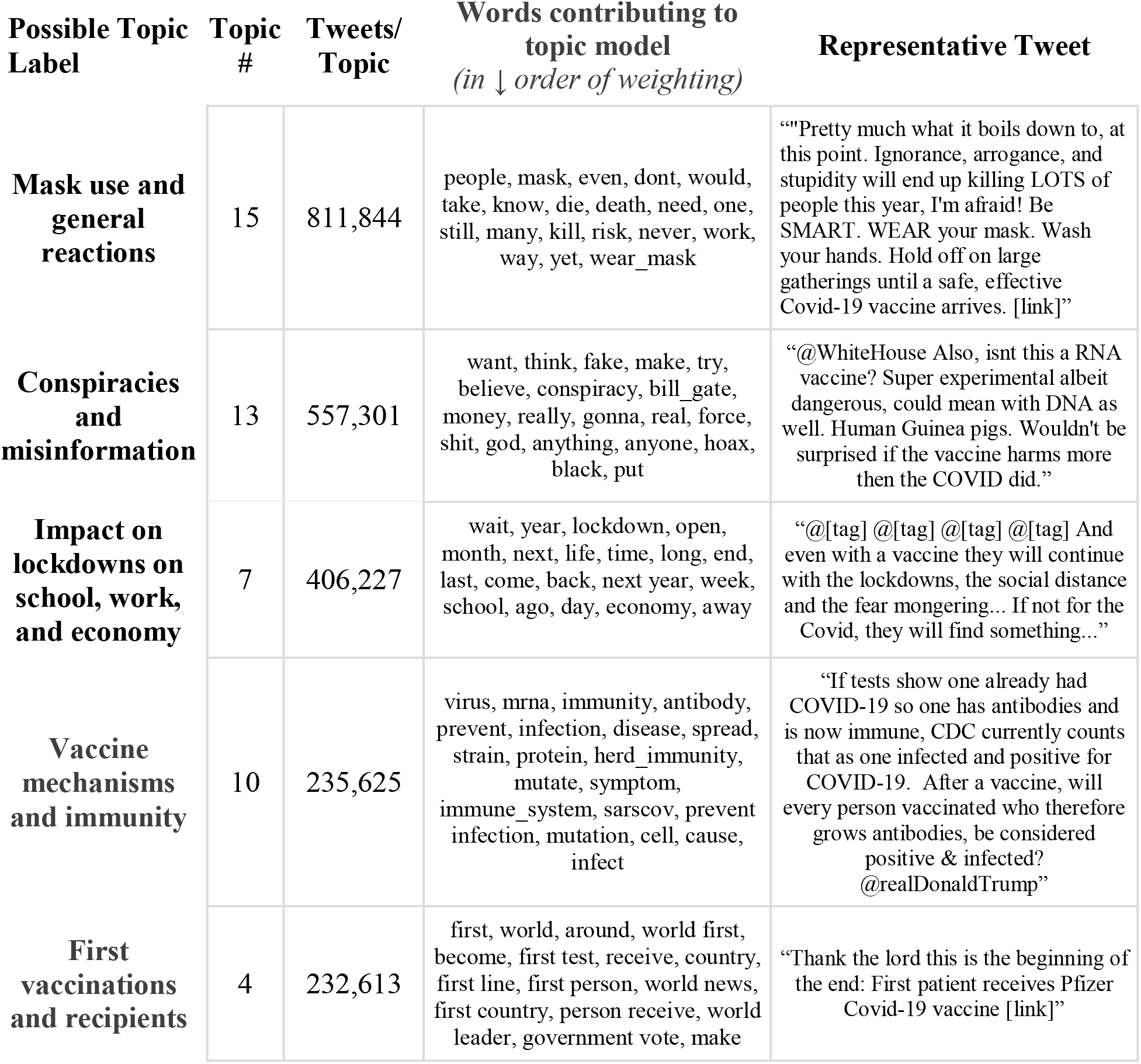
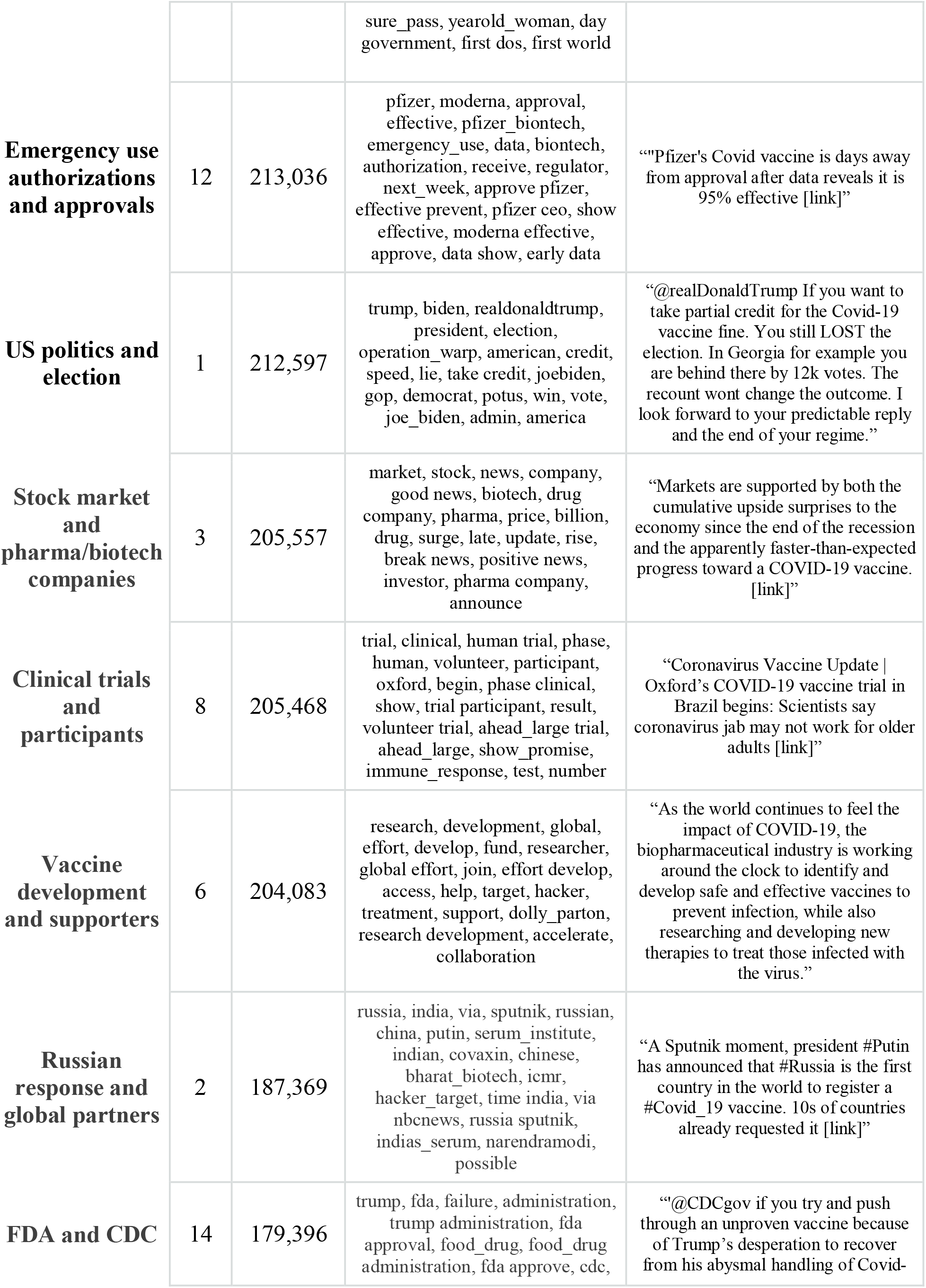
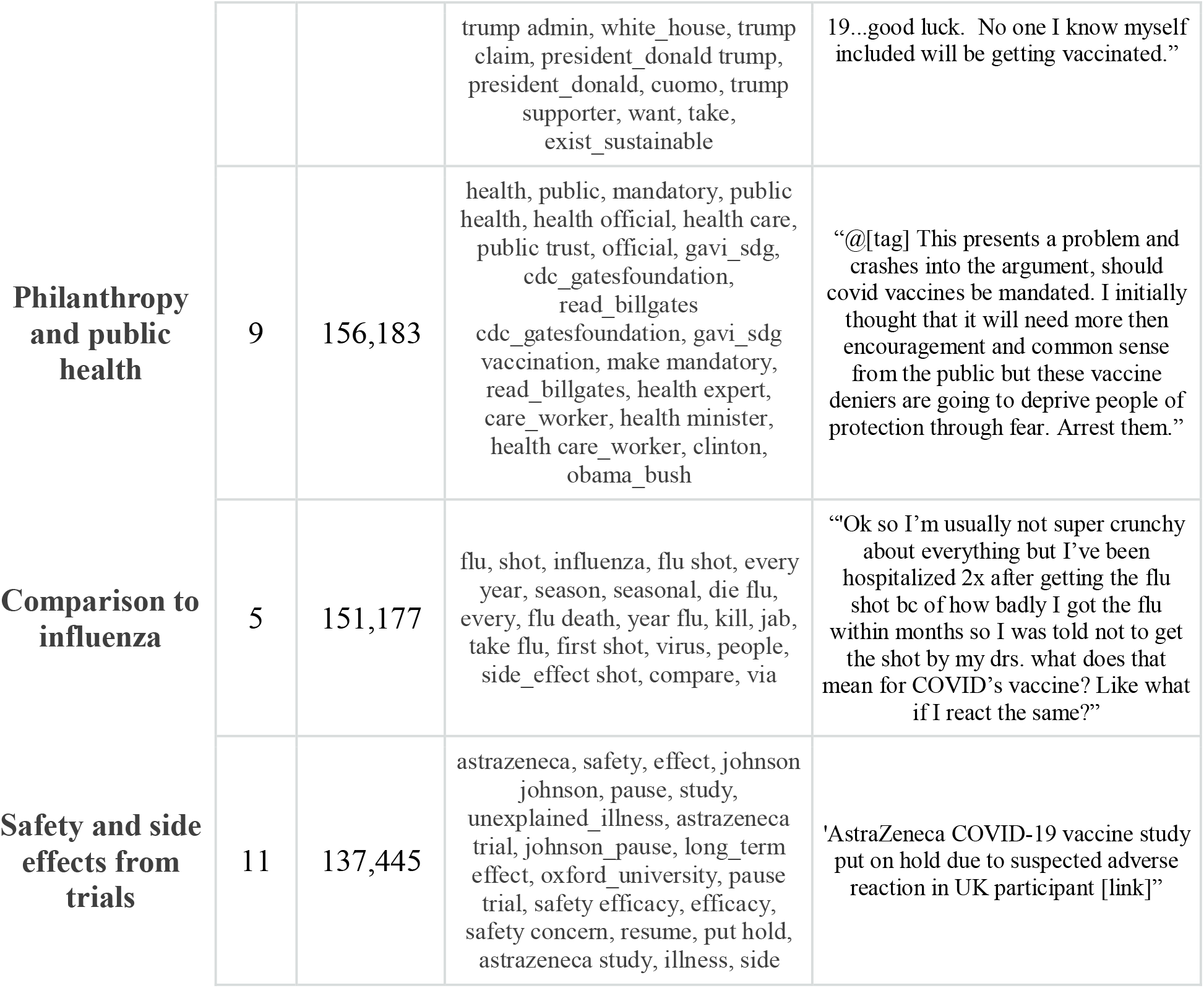
Topic clusters identified by topic modeling. Words contributing to the model are shown in decreasing order of weighting. The topics are labeled manually based on these words.

**Figure 4.**
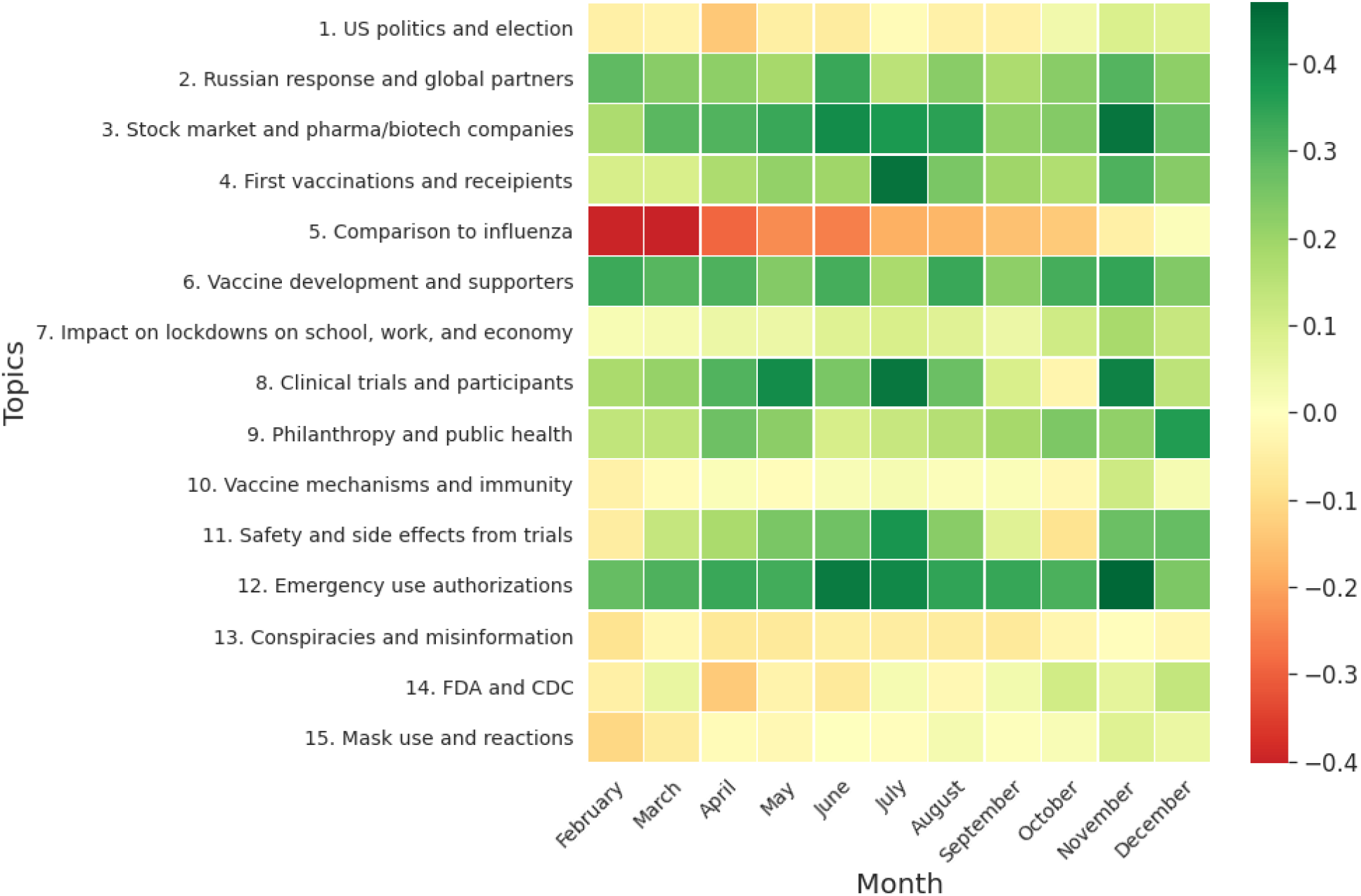
Heat map showing mean sentiment by month for each topic. Note that a tweet can include multiple topics.

## Discussion

Twitter is a rich medium that can serve as both thermometer and thermostat for the COVID-19 vaccine, which is a crucial public health strategy to combat the pandemic. It can provide insight into public perception of a COVID-19 vaccine, but can also be used to understand and combat knowledge deficits and vaccine hesitancy through information and education^41,42^. A majority (59%) of US Twitter users regularly obtain news on Twitter, proportionally more than any other social media platform^43^. We analyzed nearly 2.4 million COVID-19 vaccine related tweets in 2020, creating a dataset that exceeded the scope of related studies^44,45^. Generally, we believe that Twitter users favored the vaccine during its development phase. Tweets with positive sentiment were more prominent than tweets with negative sentiment and trust emerged as the predominant emotion. However, there were periods of time (usually linked to events in the public news cycle), demographic subgroups, and topic clusters that had more prominent negative sentiment and emotion.

Organizational accounts were significantly more positive, exhibiting more anticipation and trust and less fear. For individuals, the gender and age distribution in our dataset parallels the reported proportional share of Twitter’s global advertising audience^46^. Women expressed more fear and less anticipation, but by the end of the study period, that gap had narrowed. Those less than 40 years old tended to express less trust and more fear, but the margin was small.

The topic most strongly associated with negative, albeit improving, sentiment was the discussion of the influenza vaccine in combination with the COVID-19 vaccine. These tweets often compared deaths and illness from both diseases or expressed general vaccine mistrust to both vaccines. Examples include: *“@[user] Only time I’ve ever had the flu is the 2 times I got flu shots. It was not a minor case either it was the full blown flu. I refuse to get another flu shot and I also will refuse the covid vaccine”* and “*Flu Virus equals Flu Vaccine. Coronavirus Equals Covid-19 Vaccine*…*Now if the Flu shot gives you the flu, the Covid-19 Shot will give you Coronavirus*….*am I in the general area of Right??”*. Notably, these users exhibiting negative sentiment about this topic were more likely to be ≥ 40 years old and female. This focused topic-demographic cluster, for example, exposes a direct opportunity for intervention to correct misinformation and mitigate vaccine hesitancy. Conversely, the emergency use authorizations of the vaccine and reports of the first vaccine recipients, which arose later in the study period, were celebrated with positive sentiment and mirror the overall increasing trend in positive sentiment and trust.

While the percent population immunity needed to achieve herd immunity (either through innate or acquired immunity) for COVID-19 is not yet known, estimates have increased from 60-70% to possibly closer to 75-85%^47,48^. Achieving herd immunity through infection would come at an untenable cost^49^, making the immunization effort critical to protect lives. Therefore, it was concerning to us that fear was a common and persistent predominant emotion in COVID-19 vaccine tweets. While the proportion of ‘trust’ tweets outpaced ‘fear’ tweets relatively early in the study period, approximately 20% of eligible tweets still expressed fear in association with the vaccine. If this fear translates into refusal to become immunized, we are not only likely to see a prolonged pandemic, but also further increases in COVID-19 related deaths as concerning virus variants take hold. As more people receive the vaccine in the future, we anticipate that sentiments will become more positive over time with increased trust and vaccine uptake, but this will need to be consistently studied especially in the context of newly approved vaccines and news events.

## Limitations

Our study was limited by several factors. First, we recognize that our dataset is not all inclusive of tweets discussing the COVID-19 vaccine. Our tweet search criterion was narrow to ensure accuracy of captured tweets for this initial work and did not include terms such as “shot(s)”, “immunization” and “inoculation.” Moreover, despite the volume of tweets analyzed, we are limited to only a relevant sample of all tweets per Twitter’s advanced search tool. Second, we used existing tools to analyze sentiments and emotion of tweets that are not specific to health care topics, which could have skewed our analysis. Third, tweets related to COVID-19 vaccination could have been flagged or removed by Twitter for containing misinformation, but we were not privy to that context to determine how that could have affected our sample. Finally, since we targeted only tweets in English and are unable to determine geographic location for users, we are limited in making conclusions about specific countries or countries where English is the not the predominant language.

## Conclusions

Leveraging 2.4 million COVID-19 vaccine related tweets in 2020, we were able to successfully explore sentiment, emotion, topics, and user demographics to elucidate important trends in public perception about the COVID-19 vaccine. Tweets were overall positive in sentiment and with growing trust. However, fear maintained as a dominant emotion raising concern regarding the willingness to receive the COVID-19 vaccine and subsets of negative sentiment emerged. Comparison to influenza and the influenza vaccine as well as discussion about conspiracy theories were important topics with negative sentiment and showed some demographic differences that could allow for informed intervention. Future work will leverage these natural language processing tools to engage in targeted messaging based on user interests and emotions.

## Supporting information

Supplement - Extra Methods, Tables, and Figures

## Data Availability

The data that support the findings of this study are available upon request. Please request from the corresponding author.

## Declarations

### Ethics approval and consent to participate

The University of Texas Southwestern Human Research Protection Program Policies, Procedures, and Guidance did not require institutional review board approval as all data were publicly available.

### Code Availability

The code that support the findings of this study is available upon request. Please request from the corresponding author.

### Authors’ contributions

Study concept and design: SNS, CUL, RJM; Data acquisition: SNS, SK; Analysis: SNS, SK, RJM; Interpretation of data: SNS, SK, CUL, RJM; Manuscript preparation: all authors. All authors read and approved the final manuscript.

## Acknowledgements

Not applicable

## References

1. Sridhar, D. & Gurdasani, D. Herd immunity by infection is not an option. Science 371, 230–231s (2021).

2. Forni, G., Mantovani, A., & COVID-19 Commission of Accademia Nazionale dei Lincei, Rome. COVID-19 vaccines: where we stand and challenges ahead. Cell Death Differ 28, 626–639 (2021).

3. Caserotti, M. et al. Associations of COVID-19 risk perception with vaccine hesitancy over time for Italian residents. Soc Sci Med 272, 113688 (2021).

4. Wong, M. C. S. et al. Acceptance of the COVID-19 vaccine based on the health belief model: A population-based survey in Hong Kong. Vaccine 39, 1148–1156 (2021).

5. Feleszko, W., Lewulis, P., Czarnecki, A. & Waszkiewicz, P. Flattening the Curve of COVID-19 Vaccine Rejection-An International Overview. Vaccines (Basel) 9, (2021).

6. Ten threats to global health in 2019. https://www.who.int/news-room/spotlight/ten-threats-to-global-health-in-2019.

7. Alley, S. J. et al. As the Pandemic Progresses, How Does Willingness to Vaccinate against COVID-19 Evolve? Int J Environ Res Public Health 18, (2021).

8. Reiter, P. L., Pennell, M. L. & Katz, M. L. Acceptability of a COVID-19 vaccine among adults in the United States: How many people would get vaccinated? Vaccine 38, 6500–6507 (2020).

9. Malik, A. A., McFadden, S. M., Elharake, J. & Omer, S. B. Determinants of COVID-19 vaccine acceptance in the US. EClinicalMedicine 26, 100495 (2020).

10. Shaw, J. et al. Assessment of U.S. health care personnel (HCP) attitudes towards COVID-19 vaccination in a large university health care system. Clin Infect Dis (2021) doi:10.1093/cid/ciab054.

11. Verger, P. et al. Attitudes of healthcare workers towards COVID-19 vaccination: a survey in France and French-speaking parts of Belgium and Canada, 2020. Euro Surveill 26, (2021).

12. Saleh, S. N., Lehmann, C. U. & Medford, R. J. Early Crowdfunding Response to the COVID-19 Pandemic: Cross-sectional Study. J Med Internet Res 23, e25429 (2021).

13. Q3 2020 Letter to Shareholders. https://s22.q4cdn.com/826641620/files/doc_financials/2020/q3/Q3-2020-Shareholder-Letter.pdf.

14. McGraw, T. Spending 2020 Together on Twitter. https://blog.twitter.com/en_us/topics/insights/2020/spending-2020-together-on-twitter.html.

15. Stieglitz, S. & Dang-Xuan, L. Emotions and Information Diffusion in Social Media— Sentiment of Microblogs and Sharing Behavior. Journal of Management Information Systems 29, 217–248 (2013).

16. Depoux, A. et al. The pandemic of social media panic travels faster than the COVID-19 outbreak. Journal of Travel Medicine 27, taaa031 (2020).

17. Sinnenberg, L. et al. Twitter as a Tool for Health Research: A Systematic Review. Am J Public Health 107, e1–e8 (2017).

18. Chang, C.-H., Monselise, M. & Yang, C. C. What Are People Concerned About During the Pandemic? Detecting Evolving Topics about COVID-19 from Twitter. J Healthc Inform Res 1–28 (2021) doi:10.1007/s41666-020-00083-3.

19. Chandrasekaran, R., Mehta, V., Valkunde, T. & Moustakas, E. Topics, Trends, and Sentiments of Tweets About the COVID-19 Pandemic: Temporal Infoveillance Study. Journal of Medical Internet Research 22, e22624 (2020).

20. Xue, J. et al. Twitter Discussions and Emotions About the COVID-19 Pandemic: Machine Learning Approach. Journal of Medical Internet Research 22, e20550 (2020).

21. Gallotti, R., Valle, F., Castaldo, N., Sacco, P. & De Domenico, M. Assessing the risks of ‘infodemics’ in response to COVID-19 epidemics. Nature Human Behaviour 4, 1285–1293 (2020).

22. Medford, R. J., Saleh, S. N., Sumarsono, A., Perl, T. M. & Lehmann, C. U. An ‘Infodemic’: Leveraging High-Volume Twitter Data to Understand Early Public Sentiment for the Coronavirus Disease 2019 Outbreak. Open Forum Infect Dis 7, ofaa258 (2020).

23. Saleh, S. N., Lehmann, C. U., McDonald, S. A., Basit, M. A. & Medford, R. J. Understanding public perception of coronavirus disease 2019 (COVID-19) social distancing on Twitter. Infect Control Hosp Epidemiol 42, 131–138 (2021).

24. Systematic scoping review on social media monitoring methods and interventions relating to vaccine hesitancy. https://www.ecdc.europa.eu/en/publications-data/systematic-scoping-review-social-media-monitoring-methods-and-interventions (2020).

25. Salathé, M. & Khandelwal, S. Assessing Vaccination Sentiments with Online Social Media: Implications for Infectious Disease Dynamics and Control. PLOS Computational Biology 7, e1002199 (2011).

26. Dunn, A. G. et al. Mapping information exposure on social media to explain differences in HPV vaccine coverage in the United States. Vaccine 35, 3033–3040 (2017).

27. Wilson, S. L. & Wiysonge, C. Social media and vaccine hesitancy. BMJ Global Health 5, e004206 (2020).

28. FDA Takes Key Action in Fight Against COVID-19 By Issuing Emergency Use Authorization for First COVID-19 Vaccine. https://www.fda.gov/news-events/press-announcements/fda-takes-key-action-fight-against-covid-19-issuing-emergency-use-authorization-first-covid-19.

29. Snscrape. (Github).

30. Frijda, N. H. The emotions. (Cambridge University Press_J; Editions de la Maison des sciences de l’homme, 1986).

31. Hutto, C. & Gilbert, E.). VADER: A Parsimonious Rule-based Model for Sentiment Analysis of Social Media Text. in (2014).

32. Loria, S. Textblob: simplified text processing. (Textblob).

33. Mohammad, S. M. & Turney, P. D. Emotions evoked by common words and phrases: using mechanical turk to create an emotion lexicon. in Proceedings of the NAACL HLT 2010 Workshop on Computational Approaches to Analysis and Generation of Emotion in Text 26–34 (Association for Computational Linguistics, 2010).

34. Gallagher, R. J., Reing, K., Kale, D. & Steeg, G. V. Anchored Correlation Explanation: Topic Modeling with Minimal Domain Knowledge. arXiv:1611.10277 [cs, math, stat] (2018).

35. Steeg, G. V. & Galstyan, A. Discovering Structure in High-Dimensional Data Through Correlation Explanation. arXiv:1406.1222 [cs, stat] (2014).

36. Wang, Z. et al. Demographic Inference and Representative Population Estimates from Multilingual Social Media Data. in The World Wide Web Conference 2056–2067 (ACM, 2019). doi:10.1145/3308558.3313684.

37. Yang, Y.-C., Al-Garadi, M. A., Love, J. S., Perrone, J. & Sarker, A. Automatic Gender Detection in Twitter Profiles for Health-related Cohort Studies. http://medrxiv.org/lookup/doi/10.1101/2021.01.06.21249350 (2021) xdoi:10.1101/2021.01.06.21249350.

38. Pfizer and Biontech announce Vaccine Candidate against Covid-19 Achieved Success in First Interim Analysis From Phase 3 Study. https://www.pfizer.com/news/press-release/press-release-detail/pfizer-and-biontech-announce-vaccine-candidate-against.

39. AJMC Staff. A Timeline of COVID-19 Developments in 2020. AJMC (2021).

40. Jackson, L. A. et al. An mRNA Vaccine against SARS-CoV-2 - Preliminary Report. N Engl J Med 383, 1920–1931 (2020).

41. Subbaraman, N. This COVID-vaccine designer is tackling vaccine hesitancy - in churches and on Twitter. Nature 590, 377 (2021).

42. Loomba, S., de Figueiredo, A., Piatek, S. J., de Graaf, K. & Larson, H. J. Measuring the impact of COVID-19 vaccine misinformation on vaccination intent in the UK and USA. Nat Hum Behav 5, 337–348 (2021).

43. Shearer, E. & Mitchell, A. News Use Across Social Media Platforms in 2020. https://www.journalism.org/2021/01/12/news-use-across-social-media-platforms-in-2020/ (2021).

44. Damiano, A. D. & Allen Catellier, J. R. A Content Analysis of Coronavirus Tweets in the United States Just Prior to the Pandemic Declaration. Cyberpsychol Behav Soc Netw 23, 889–893 (2020).

45. Jang, H., Rempel, E., Roth, D., Carenini, G. & Janjua, N. Z. Tracking COVID-19 Discourse on Twitter in North America: Infodemiology Study Using Topic Modeling and Aspect-Based Sentiment Analysis. J Med Internet Res 23, e25431 (2021).

46. Kemp, S. Digital 2020 Global Digital Overview. https://p.widencdn.net/1zybur/Digital2020Global_Report_en (2020).

47. Fontanet, A. & Cauchemez, S. COVID-19 herd immunity: where are we? Nat Rev Immunol 20, 583–584 (2020).

48. McNeil Jr., D.G. https://www.nytimes.com/2020/12/24/health/herd-immunity-covid-coronavirus.html. The New York Times (2021).

49. Randolph, H. E. & Barreiro, L. B. Herd Immunity: Understanding COVID-19. Immunity 52, 737–741 (2020).

