## Supplement - Extra Methods, Tables, and Figures for "Public Perception of COVID-19 Vaccines through Analysis of Twitter Content and Users"

### Appendix A.

#### Methods: data processing, transformation, and exploration.

For sentiment and emotion analyses, we limited preprocessing content to maximize context and expression (including punctuation, capitalization, emojis, and emoticons). We only removed any html syntax, hyperlinks, and user mentions. For topic modeling, we further removed punctuation, converted the tweet plain text to lowercase, changed words to their root forms (e.g., ‘viruses’ to ‘virus’ or ‘went’ to ‘go’) using the WordNetLemmatizer module of the *NLTK* library in Python(1), and removed stop words (frequently used words with little semantic meaning, such as ‘of’, ‘it’, and ‘is’). For additional dimensionality reduction, we created a list of n-grams (one-word and two-word terms or unigrams and bigrams, respectively) from tweets and removed all extremely low (present in less than 10 total tweets) and high frequency (present in over 25% of tweets) terms. These steps decreased the dictionary of terms from 746,045 to 59,635.

**Table S1.** Metadata associated with tweets

| url | date | content |
| --- | --- | --- |
| renderedContent | id | user |
| outlinks | tcooutlinks | replyCount |
| retweetCount | likeCount | quoteCount |
| conversationId | lang | source |
| sourceUrl | sourceLabel | Media |
| retweetedTweet | quotedTweet | mentionedUsers |

**Table S2.** Metadata associated with users

| username | displayname | Id |
| --- | --- | --- |
| description | rawDescription | descriptionUrls |
| verified | created | followersCount |
| friendsCount | statusesCount | favouritesCount |
| listedCount | mediaCount | location |
| protected | linkUrl | profileImageUrl |
| profileBannerUrl | url |  |

**Figure S1.** Number of COVID-19 vaccine related tweets over time by day.

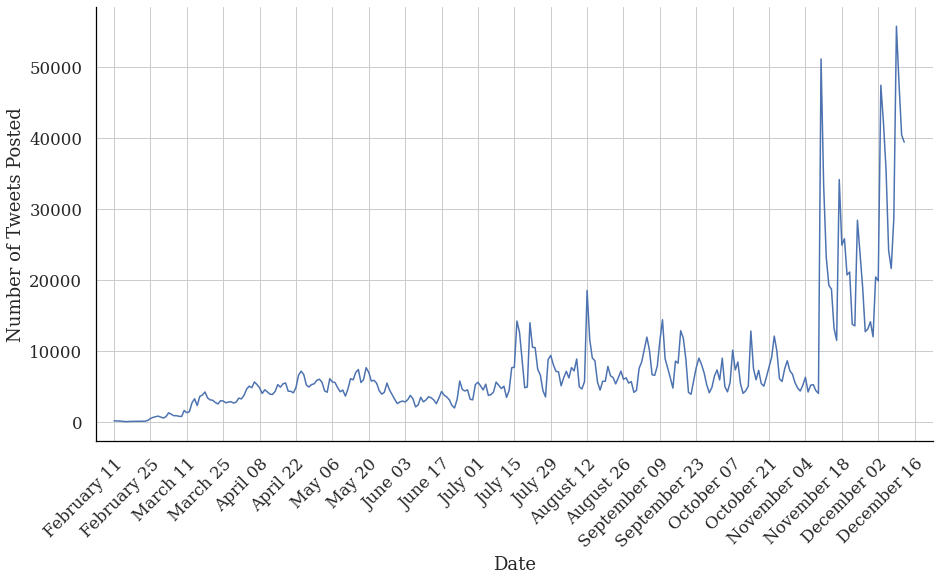

**Figure S2.** Mean subjectivity by day with 0 as fully objective and 1 as fully subjective.

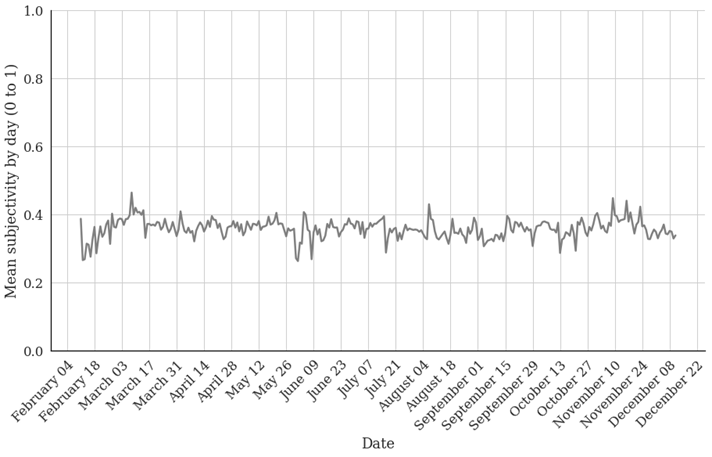

**Figure S3.** Proportion of topics by month containing each topic.

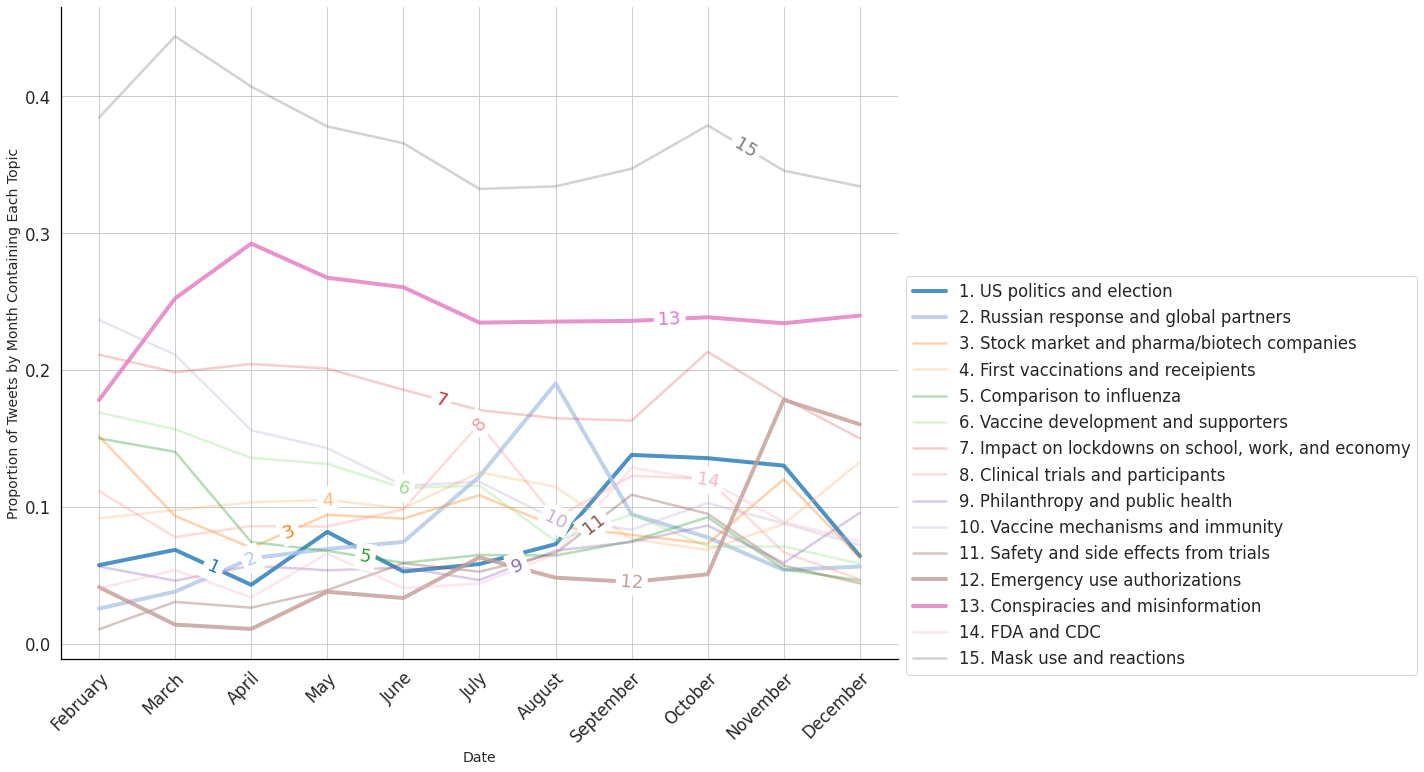
